# Ambient air pollution and COVID-19 in National Capital Territory of Delhi, India: a time-series evidence

**DOI:** 10.1101/2021.06.04.21258376

**Authors:** Abhishek Singh

**Affiliations:** Department of Mathematics and Scientific Computing, National Institute of Technology Hamirpur, Anu Road, Hamirpur, Himachal Pradesh, 177005

**Keywords:** COVID-19, air pollution, population mobility, generalized additive model, India

## Abstract

**Objectives:** This study aimed to explore the short-term health effects of ambient air pollutants *PM*_2.5_, *PM*_10_, *SO*_2_, *NO*_2_, *O*_3_, and *CO* on COVID-19 daily new cases and COVID-19 daily new deaths.

**Study design:** A time-series design used in this study. Data were obtained from 1 April 2020 to 31 December 2020 in the National Capital Territory (NCT) of Delhi, India.

**Methods:** The generalized additive models (GAMs) were applied to explore the associations of six air pollutants with COVID-19 daily new cases and COVID-19 daily new deaths. We also conducted sensitivity analysis using the population mobility variable in terms of lockdowns.

**Results:** The GAMs revealed statistically significant associations of ambient air pollutants with COVID-19 daily new cases and COVID-19 daily new deaths. Besides, in sensitivity analysis after controlling for the population mobility, these associations became more prominent.

**Conclusions:** These findings suggest that governments need to give greater considerations to regions with higher concentrations of *PM*_2.5_, *PM*_10_, *SO*_2_, *NO*_2_, *O*_3_, and *CO*, since these areas may experience a more serious COVID-19 pandemic or, in general, any respiratory disease.

## Introduction

The world health organization (WHO) declared the coronavirus disease 2019 (COVID-19) a global pandemic first detected in Wuhan, China in December 2019 (Lu et al., 2020; Wang et al., 2020; Xu et al., 2020; Zhu et al., 2020). COVID-19 is a highly transmissible and fatal disease induced by the severe acute respiratory syndrome coronavirus 2 (SARS-CoV-2) (Dong et al., 2020; Xie and Zhu, 2020; Zhou et al., 2020). Usually, maximum COVID-19 infected patients showed mild to moderate symptoms including sore throat, fever, shortness of breath, dry cough, and loss of smell and taste with some severe patients having pneumonia, acute respiratory distress syndrome (ARDS), kidney failure, and even death (Ali and Alharbi, 2020; Chen et al., 2020; Menni et al., 2020; Sohrabi et al., 2020).

Previous studies argued that ambient air pollutants are risk factors associated with respiratory infection and respiratory infection-induced death (Atkinson et al., 2014; Balakrishnan et al., 2019; Cai et al., 2007; Cui et al., 2003; Horne et al., 2018; Siddique et al., 2011; Xie et al., 2019; Xu et al., 2016). COVID-19 is also a respiratory disease and SARS-CoV-2 could remain effective and infectious in aerosols for a number of days (Doremalen et al., 2020). Many studies have demonstrated the interrelation of short-term and chronic exposure to ambient air pollution with COVID-19 infection (Fattorini and Regoli, 2020; Frontera et al., 2020; Muhammad et al., 2020; Ogen, 2020; Zhu et al., 2020). Besides, the literature also demonstrated the association of temperature and humidity with COVID-19 infection (Badr et al., 2020; Y. Wu et al., 2020; Xie and Zhu, 2020). Also, many previous studies reported that interpersonal contact could increase the transmission of SARS-CoV-2 infection (Auler et al., 2020; Bashir et al., 2020; Briz-Redón and Serrano-Aroca, 2020; Chan et al., 2020; Ghinai et al., 2020; Li et al., 2020). It is also established that population mobility has a significant effect on the spread of COVID-19 infection (Badr et al., 2020; Kraemer et al., 2020; Wang et al., 2020).

Recently effect of ambient air pollution on COVID-19 infection has been reported extensively, and only few studies have been conducted to investigate the short-term effect of air pollution on COVID-19 deaths. Besides, to the best of our knowledge, not a single study has been taken into account the population mobility variable in terms of lockdowns to see the effect of ambient air pollution on COVID-19 daily new cases and COVID-19 daily new deaths. The present study aimed to estimate the short-term effects of *PM*_2.5_, *PM*_10_, *SO*_2_, *NO*_2_, *O*_3_, and *CO* on COVID-19 daily new cases and COVID-19 daily new deaths after controlling for the meteorological factors and population mobility in terms of lockdowns in India.

## Methods

The current study was conducted in the National Capital Territory (NCT) of Delhi (28°36′36″*N* 77°13′ 48″ *E*). The NCT of Delhi is the largest city in India. It is a union territory and contains the capital of India, New Delhi. The complete area of the NCT of Delhi is about 1484.0 km^2^ with 16.8 million inhabitants, representing 1.4% of India’s total population (Census of India 2011). In 2018, it was listed as one of the most polluted cities on earth by the WHO global air pollution database released in Geneva (Times of India 2018).

### Data source

We obtained data, including daily new cases and daily new deaths of COVID-19 from the Ministry of Health and Family Welfare, Government of India (https://www.mohfw.gov.in) from 1 April 2020 to 31 December 2020. Daily air pollution and meteorological data were extracted from the database of the Central Pollution Control Board, Ministry of Environment, Forest and Climate Change, Government of India (https://cpcb.nic.in). Daily air pollution data include particles with diameters *≤* 2.5*µm* (*PM*_2.5_), particles with diameters *≤* 10*µm* (*PM*_10_), sulfur dioxide (*SO*_2_), nitrogen dioxide (*NO*_2_), Ozone (*O*_3_), and carbon monoxide (*CO*). We evaluated three meteorological variables: daily mean temperature, relative humidity, and wind speed during the study period. We averaged the daily concentrations of air pollutants from the 38 monitoring stations and daily meteorological data from 4 monitoring stations in the NCT of Delhi as the proxy for the common exposure for all residents. Data on population mobility in terms of lockdowns also obtained from 1 April 2020 to 31 December 2020 from the Ministry of Home Affairs, Government of India (https://www.mha.gov.in).

### Statistical analysis

Spearman’s correlation coefficients were used to assess the interrelations between air pollutants and meteorological variables during the study period. COVID-19 daily new cases and COVID-19 daily new deaths were linked with air pollution concentrations by date and, therefore, can be analyzed with a time-series study design. The time-invariant confounders at the population level are automatically controlled in the time-series study design. (Chen et al., 2018).

As demonstrated in many previous time-series studies of air pollution epidemiology, it is an optimal practice to use a moving average method to obtain the cumulative lag effect of ambient air pollution (Jiang and Xu, 2021; Kan et al., 2008; Li et al., 2018; Phosri et al., 2019; Wang et al., 2020; Xie et al., 2019; Yang et al., 2020; Zhang et al., 2015). As daily new cases and daily new deaths of COVID-19 approximately followed a negative binomial distribution, we used the generalized additive model with natural cubic spline smoothers for meteorological variables to investigate the cumulative lag effects (lag0-7, lag0-14, and lag0-21) of each air pollutants on COVID-19 daily new cases and COVID-19 daily new deaths. We analyzed the associations of six air pollutants with daily new cases and daily new deaths of COVID-19 in six separate models to reduce the problem of collinearity as some of these ambient air pollutants were extremely correlated. To control for confounding of meteorological variables, we used 6 df for the smoothing of daily mean temperature and 3 df for the smoothing of relative humidity, and wind speed (Cao et al., 2012; Chen et al., 2010; Phosri et al., 2019). The following equation was used to fit the generalized additive models:

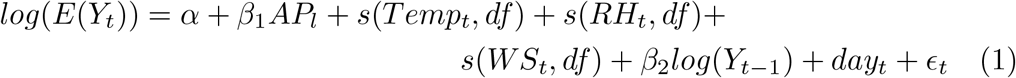

Here, *E*(*Y*_*t*_) specifies the estimated number of COVID-19 daily new cases and COVID-19 daily new deaths at day *t. α* is the intercept; *β* represents the regression coefficient; *AP*_*l*_ indicates the *l−day* moving average concentrations of air pollutants *PM*_2.5_, *PM*_10_, *SO*_2_, *NO*_2_, *O*_3_, and *CO* at lag 0− *l. s*(*Temp*_*t*_, *df*), *s*(*RH*_*t*_, *df*), and *s*(*WS*_*t*_, *df*) are the natural cubic spline functions for temperature, humidity, and wind speed, respectively. *log*(*Y*_*t −* 1_) denotes the log-transformed of COVID-19 daily new cases and COVID-19 daily new deaths reported on day *t−* 1 to account for the potential autocorrelation present in data. We also included fixed effects at day *t* (*day*_*t*_) to control unobserved components affecting the National Capital Territory of Delhi, India.

### Sensitivity Analysis

Many previous studies had found that the transmission of infectious disease affected by population mobility (Bajardi et al., 2011; Cassels, 2020; Hufnagel et al., 2004). Most part of the world was in complete lockdown to mitigate and contain the transmission of highly infectious COVID-19 disease. The complete lockdown in India including the NCT of Delhi was imposed from 25 March 2020 to 31 May 2020 by the Ministry of Home Affairs, Government of India to control the spread of the COVID-19. After remaining in the complete lockdown for over two months, the country was reopened in six different phases from 1 June 2020 to 30 November 2020. These phases were put into effect in terms of Unlock-1.0 (1 June 2020 -30 June 2020), Unlock-2.0 (1 July 2020 -31 July 2020), Unlock-3.0 (1 August 2020 -31 August 2020), Unlock-4.0 (1 September 2020 -30 September 2020), Unlock-5.0 (1 October 2020 -31 October 2020), and finally Unlock-6.0 (1 November 2020 -30 November 2020). The terms and conditions for each Unlock’s was relaxed gradually to get the full population mobility by the Unlock-6 (1 November 2020 -30 November 2020) and it was further extended up to 31 December 2020. Thus in sensitivity analysis, the lockdown variable has been considered as the categorical proxy variable for population mobility indicating ‘0’ as no population mobility and ‘6’ as full population mobility.

The statistical analyses in this study were conducted using the “mgcv” package (version 1.8.26) (Wood, 2011) in R statistical software (version 3.5.2) (R Core Team, 2018). The statistical tests were two-sided, and *p <* 0.05, *p <* 0.01, and *p <* 0.001 were considered statistically significant.

## Results

### Descriptive analysis of data on COVID-19 daily new cases and COVID-19 daily new deaths, air pollutants, and meteorological factors

A total of 627,113 confirmed cases of COVID-19 with an average of 2,280 and 10,796 COVID-19 deaths with an average of 39 were recorded from 1 April 2020 to 31 December 2021 in National Capital Territory of Delhi, India. The daily mean concentrations of air pollutants were 88.3 *µg/m*^3^ for *PM*_2.5_, 175.3 *µg/m*^3^ for *PM*_10_, 13.3 *µg/m*^3^ for *SO*_2_, 36.5 *µg/m*^3^ for *NO*_2_, 35.2 *µg/m*^3^ for *O*_3_, and 1.2 *µg/m*^3^ for CO. The average daily mean temperature, relative humidity, and wind speed were 27.5 °*C*, 57.8%, and 1.1%, respectively (see Table S1 in the supplementary material).

Spearman’s correlation coefficients between air pollutants and meteorological variables have been presented in Table 1. Air pollutants had a significant correlation with each other. *PM*_2.5_ had a strong correlation (0.96) with *PM*_10_, followed by the correlation (0.89) between *NO*_2_ and CO. All ambient air pollutants were correlated with mean temperature, relative humidity, and wind speed except for CO and relative humidity. The air pollutants *PM*_2.5_, *PM*_10_, *SO*_2_, and *NO*_2_, were inversely correlated with mean temperature, relative humidity, and wind speed. *O*_3_ was positively correlated with mean temperature and wind speed. However, it was inversely correlated with relative humidity. Besides, CO was also inversely correlated with mean temperature and wind speed. The results suggested that it was necessary to adjust the influence of coexisting meteorological factors in analyzing the associations of air pollutants with daily new cases and daily new deaths of COVID-19.

**Table 1:** Spearman’s correlation coefficients between air pollutants and meteorological variables in the National Capital Territory of Delhi, India during the study period (1 April 2020 through 31 December 2021).

| | $PM_{2.5}$ | $PM_{10}$ | $SO_2$ | $NO_2$ | $O_3$ | CO | Mean temperature | Relative humidity | Wind speed |
| --- | --- | --- | --- | --- | --- | --- | --- | --- | --- |
| $PM_{2.5}$ | 1.00 | | | | | | | | |
| $PM_{10}$ | 0.96* | 1.00 | | | | | | | |
| $SO_2$ | 0.70* | 0.70* | 1.00 | | | | | | |
| $NO_2$ | 0.84* | 0.82* | 0.52* | 1.00 | | | | | |
| $O_3$ | 0.16* | 0.18* | 0.52* | -0.15* | 1.00 | | | | |
| CO | 0.82* | 0.78* | 0.48* | 0.89* | -0.18* | 1.00 |  |  |  |
| Mean temperature | -0.55* | -0.48* | -0.36* | -0.52* | 0.11 | -0.48* | 1.00 |  |  |
| Relative humidity | -0.32* | -0.40* | -0.66* | -0.10 | -0.73* | 0.00 | -0.05 | 1.00 |  |
| Wind speed | -0.59* | -0.54* | -0.43* | -0.73* | 0.07 | -0.68* | 0.33* | 0.05 | 1.00 |

Note: \* $p < 0.05$ .

### Effects of ambient air pollution on COVID-19 daily new cases

The effects of ambient air pollution on COVID-19 daily new cases in NCT of Delhi were statistically significant at some lag days only (Table 2). After controlling the effects of possible confounders, we found statistically significant associations of *PM*_2.5_, *PM*_10_, *SO*_2_, and *NO*_2_ with COVID-19 daily new cases. The associations remained statistically significant only in the moving average concentrations of 14 days and 21 days. For the moving average concentrations of 14 days, the associations of *PM*_2.5_, *PM*_10_, and *NO*_2_ were positive and significant with COVID-19 daily new cases. A 10− *µg/m*^3^ increase in concentrations (lag0-14 days) in *PM*_2.5_ was associated with a 1.46% (95% CI: 0.22%, 2.70%) increase in COVID-19 daily new cases. Similarly, A 10*− µg/m*^3^ increase in concentrations (lag0-14 days) in *PM*_10_ and *NO*_2_ was associated with a 1.06% (95% CI: 0.24%, 1.89%) and 8.97% (95% CI: 3.14%, 14.80%) increase in COVID-19 daily new cases, respectively. In moving average concentrations of 21 days, associations of *PM*_10_, *SO*_2_, and *NO*_2_ were positive and significant with COVID-19 daily new cases. A 10*− µg/m*^3^ increase in concentrations (lag0-21 days) in *PM*_10_, *SO*_2_, and *NO*_2_ was associated with a 1.03% (95% CI: 0.15%, 1.90%), 37.23% (95% CI: 10.62%, 63.85%), and 8.41% (95% CI: 2.38%, 14.45%) increase in COVID-19 daily new cases, respectively.

**Table 2:** Percent change (means and 95% confidence intervals) in COVID-19 daily new cases associated with a 10 *—µg/m*^3^ increase in *PM*_2.5_, *PM*_10_, *SO*_2_, *NO*_2_, and *O*_3_ and 1−*µg/m*^3^ increase in *CO* using different lag days in single pollutant models.

| lag | $PM_{2.5}$ | $PM_{10}$ | $SO_2$ | $NO_2$ | $O_3$ | $CO$ |
| --- | --- | --- | --- | --- | --- | --- |
| lag0-7 | 0.61 (-0.42, 1.64) | 0.44 (-0.25, 1.14) | 10.89 (-13.53, 35.30) | 4.69 (-0.68, 10.05) | 2.19 (-4.20, 8.58) | 4.18 (-10.07, 18.43) |
| lag0-14 | 1.46* (0.22, 2.70) | 1.06* (0.24, 1.89) | 26.13 (-0.59, 52.85) | 8.97** (3.14, 14.80) | 3.58 (-3.90, 11.06) | 18.28 (0.33, 36.23) |
| lag0-21 | 1.22 (-0.10, 2.53) | 1.03* (0.15, 1.90) | 37.23** (10.62, 63.85) | 8.41** (2.38, 14.45) | 6.03 (-1.26, 13.32) | 17.14 (-2.12, 36.41) |

Note: \* $p < 0.05$ and \*\* $p < 0.01$ . Abbreviations: lag0-7: the moving average concentrations of 7 days, lag0-14: the moving average concentrations of 14 days, and lag0-21: the moving average concentrations of 21 days. The model covariates include mean temperature, relative humidity, wind speed, log-transformed daily new cases of COVID-19 on day $t - 1$ , and day fixed effects.

The associations of *PM*_2.5_, *PM*_10_, *SO*_2_, *NO*_2_, *O*_3_, and *CO* with daily new COVID-19 cases were usually positive, however, statistically insignificant in the ambient air pollution moving average concentrations of 7 days. For example, a 10*− µg/m*^3^ increase in concentrations (lag0-7 days) of *PM*_2.5_, *PM*_10_, *SO*_2_, *NO*_2_, and *O*_3_ was associated with percent change of 0.61% (95% CI: -0.42%, 1.64%), 0.44% (95% CI: -0.25%, 1.14%), 10.89% (95% CI: -13.53%, 35.30%), 4.69% (95% CI: -0.68%, 10.05%), and 2.19% (95% CI: -4.20%, 8.58%) in COVID-19 daily new cases, respectively. Similarly, a 1*− µg/m*^3^ increase in concentrations (lag0-7 days) of *CO* was associated with percent change of 4.18% (95% CI: -10.07%, 18.43%) in COVID-19 daily new cases.

### Effects of ambient air pollution on COVID-19 daily new deaths

The effects of all ambient air pollution on COVID-19 daily new deaths in NCT of Delhi were statistically significant for lag0-14 and lag0-21 days (Table 3). For the moving average concentrations of 14 days, the associations of *PM*_2.5_, *PM*_10_, *SO*_2_, *NO*_2_, and *O*_3_ were positive and significant with COVID-19 daily new deaths. A 10−*µg/m*^3^ increase in concentrations (lag0-14 days) in *PM*_2.5_ was associated with a 5.13% (95% CI: 2.71%, 7.54%) increase in COVID-19 daily new deaths. Similarly, A 10*− µg/m*^3^ increase in concentrations (lag0-14 days) in *PM*_10_, *SO*_2_, *NO*_2_, and *O*_3_ was associated with a 3.86% (95% CI: 2.34%, 5.39%), 134.88% (95% CI: 84.03%, 185.73%), 33.56% (95% CI: 21.78%, 45.34%), and 43.54% (95% CI: 30.17%, 56.91%) increase in COVID-19 daily new deaths, respectively. A 1*− µg/m*^3^ increase in concentrations (lag0-14 days) in *CO* was associated with a 112.19% (95% CI: 73.81%, 150.57%) increase in COVID-19 daily new deaths. In moving average concentrations of 21 days, the associations of *PM*_2.5_, *PM*_10_, *SO*_2_, *NO*_2_, *O*_3_, and *CO* were positive and significant with COVID-19 daily new deaths. A 10−*µg/m*^3^ increase in concentrations (lag0-21 days) in *PM*_2.5_, *PM*_10_, *SO*_2_, *NO*_2_, and *O*_3_ was associated with a 8.91% (95% CI: 6.03%, 11.80%), 6.77% (95% CI: 4.97%, 8.57%), 213.57% (95% CI: 158.05%, 269.10%), 48.49% (95% CI: 39.94%, 62.04%), and 52.87% (95% CI: 42.50%, 73.24%) increase in COVID-19 daily new deaths, respectively. Similarly, a 1−*µg/m*^3^ increase in concentrations (lag0-21 days) in *CO* was associated with a 236.45% (95% CI: 188.77%, 284.13%) increase in COVID-19 daily new deaths.

**Table 3:** Percent change (means and 95% confidence intervals) in COVID-19 daily new deaths associated with a 10 *— µg/m*^3^ increase in *PM*_2.5_, *PM*_10_, *SO*_2_, *NO*_2_, and *O*_3_ and 1−*µg/m*^3^ increase in *CO* using different lag days in single pollutant models.

| lag | $PM_{2.5}$ | $PM_{10}$ | $SO_2$ | $NO_2$ | $O_3$ | $CO$ |
| --- | --- | --- | --- | --- | --- | --- |
| lag0-7 | 1.58 (-0.37, 3.52) | 1.33* (0.07, 2.60) | 62.66** (19.30, 106.02) | 17.32** (6.77, 27.88) | **27.09 (16.03, 38.15)** | 18.81 (-10.85, 48.47) |
| lag0-14 | **5.13 (2.71, 7.54)** | **3.86 (2.34, 5.39)** | **134.88 (84.03, 185.73)** | **33.56 (21.78, 45.34)** | **43.54 (30.17, 56.91)** | **112.19 (73.81, 150.57)** |
| lag0-21 | **8.91 (6.03, 11.80)** | **6.77 (4.97, 8.57)** | **213.57 (158.05, 269.10)** | **48.49 (34.94, 62.04)** | **57.87 (42.50, 73.24)** | **236.45 (188.77, 284.13)** |

Note: \* $p < 0.05$ , \*\* $p < 0.01$ , and $p < 0.001$ are highlighted in bold. Abbreviations: lag0-7: the moving average concentrations of 7 days, lag0-14: the moving average concentrations of 14 days, and lag0-21: the moving average concentrations of 21 days. The model covariates include mean temperature, relative humidity, wind speed, log-transformed daily new cases of COVID-19 on day $t - 1$ , and day fixed effects.

The associations of *PM*_2.5_ and *CO* with daily new COVID-19 deaths were positive and statistically insignificant in the ambient air pollution moving average concentrations of 7 days. For example, a 10− *µg/m*^3^ increase in concentrations (lag0-7 days) of *PM*_2.5_ was associated with the percent change of 1.58% (95% CI: -0.37%, 3.52%) in COVID-19 daily new deaths. Similarly, a 1*− µg/m*^3^ increase in concentrations (lag0-7 days) of *CO* was associated with the percent change of 18.81% (95% CI: -10.85%, 48.47%) in COVID-19 daily new deaths.

### Sensitivity analysis

Population mobility variable was adjusted in each model to examine the change in the effects of air pollutants on COVID-19 daily new cases and COVID-19 daily new deaths. The results revealed that a 10− *µg/m*^3^ increase in concentrations (lag0-14 days) in *PM*_2.5_ was associated with a 1.96% (95% CI: 0.36%, 3.56%) increase in COVID-19 daily new cases. Similarly, a 10-*µg/m*^3^ increase in concentrations (lag0-14 days) in *PM*_10_ and *NO*_2_ was associated with a 1.20% (95% CI: 0.18%, 2.23%) and 10.29% (95% CI: 3.20%, 17.36%) increase in COVID-19 daily new cases, respectively. Further, a 1*− µg/m*^3^ increase in concentrations (lag0-14 days) in *CO* was associated with a 27.43% (95% CI: 4.26%, 50.61%) increase in COVID-19 daily new cases. In moving average concentrations of 21 days, the effects of all ambient air pollutants were positive and significant for COVID-19 daily new cases (Fig. 1 and Table S2 in the supplementary material).

**Figure 1:**
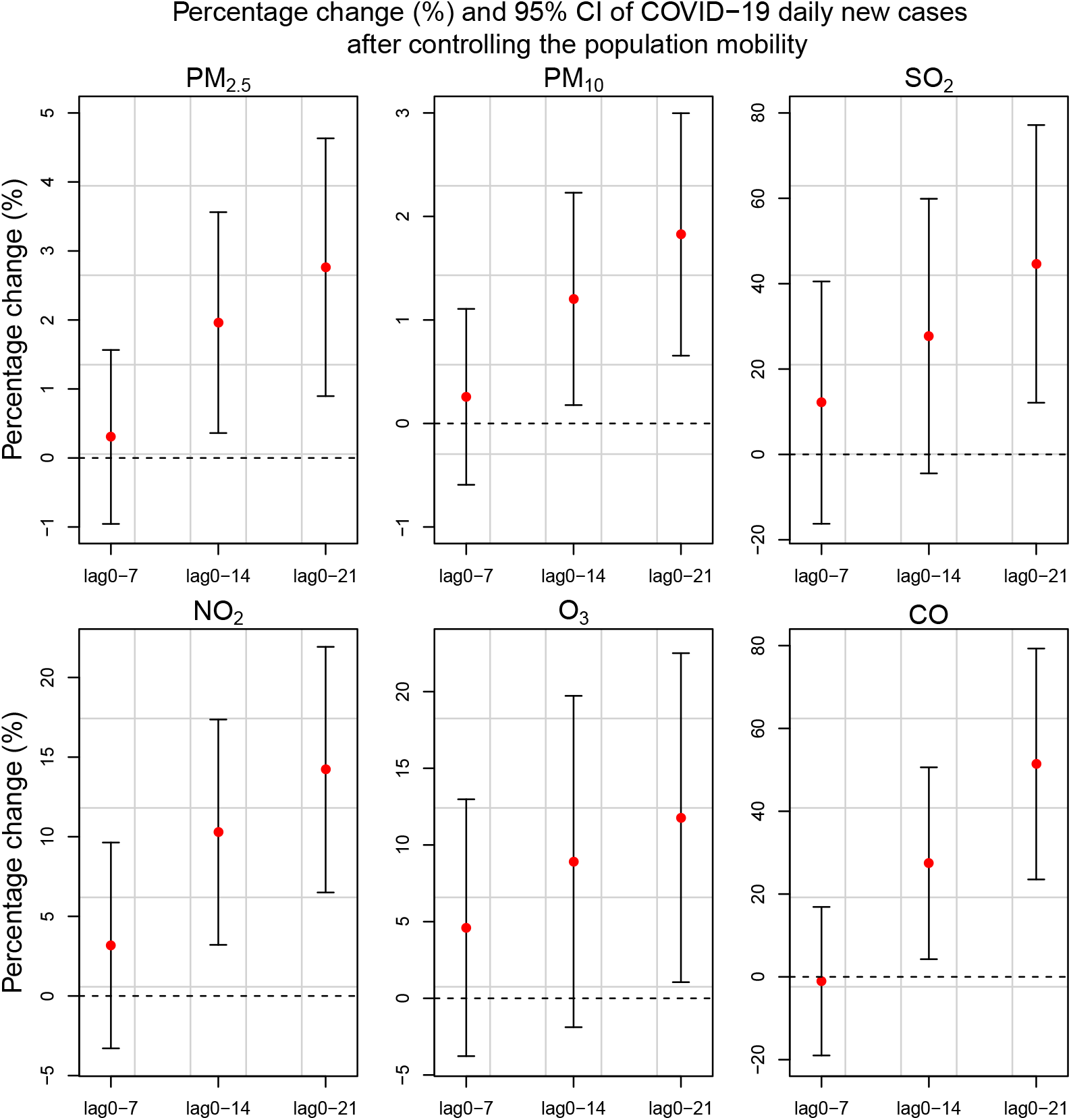
Sensitivity analysis: percentage change (%) and 95% CI of COVID-19 daily new cases associated with ambient air pollution

For the moving average concentrations of 7 days, the associations of *NO*_2_ and *O*_3_ with COVID-19 daily new deaths were positive and significant. A 10-*µg/m*^3^ increase in concentrations (lag0-7 days) in *NO*_2_ and *O*_3_ was associated with a 9.33% (95% CI: 1.07%, 17.58%) and 27.03% (95% CI: 14.23%, 39.83%) increase in COVID-19 daily new deaths. In moving average concentrations of 14 and 21 days, the effects of all ambient air pollutants were positive and significant for COVID-19 daily new deaths (Fig. 2 and Table S3 in the supplementary material).

**Figure 2:**
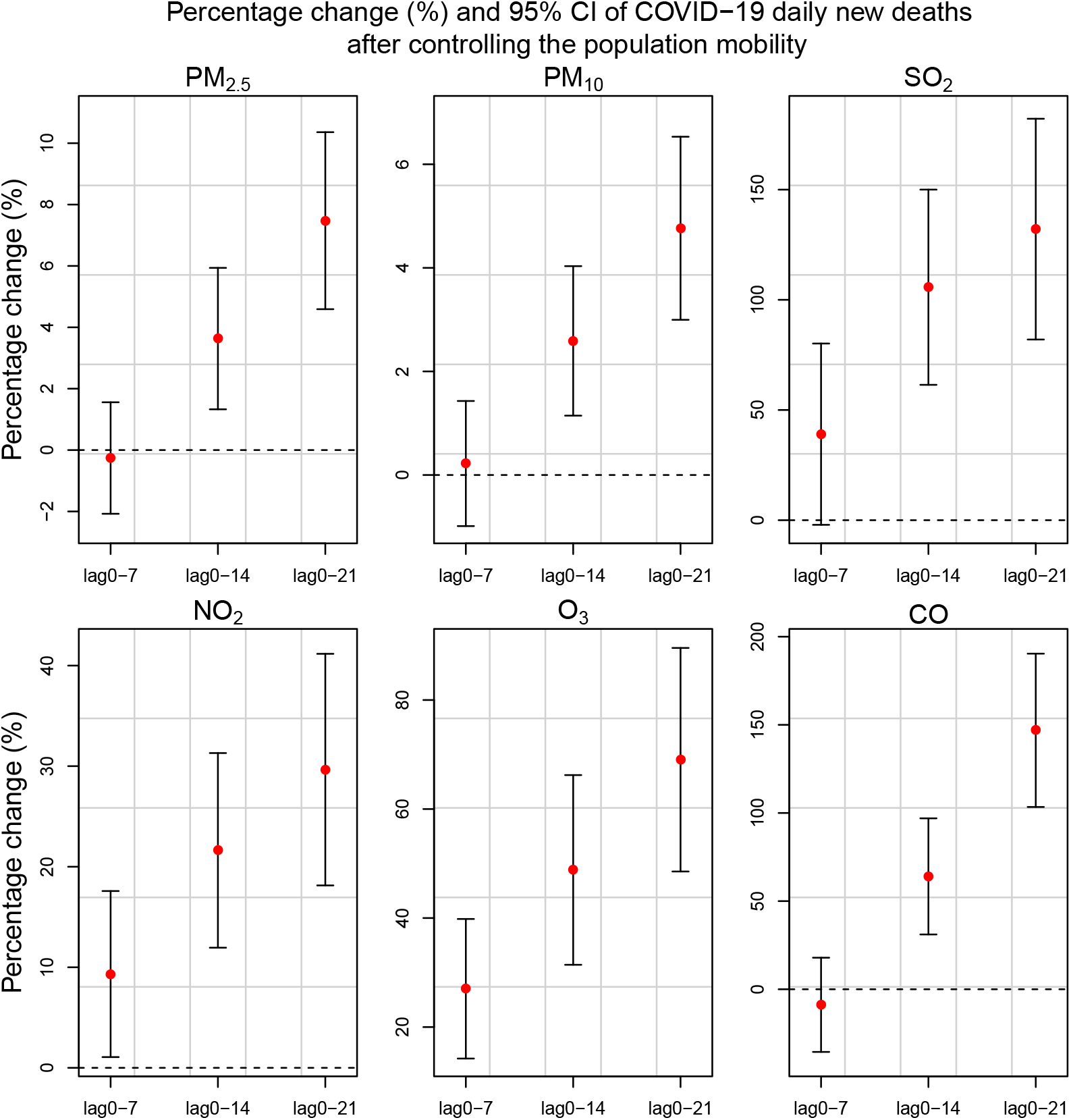
Sensitivity analysis: percentage change (%) and 95% CI of COVID-19 daily new deaths associated with ambient air pollution

## Discussion

As far as we know, this is the pioneer study to explore the effects of air pollution on COVID-19 daily new cases and COVID-19 daily new deaths by adjusting for the population mobility in terms of lockdowns, employing time-series study design. This study used generalized additive models with a negative binomial distribution to examine the effects of ambient air pollution on COVID-19 daily new cases and COVID-19 daily new deaths. The nine months data from the National Capital Territory of Delhi, India, showed the significant associations between daily concentrations of ambient air pollution (*PM*_2.5_, *PM*_10_, *SO*_2_, *NO*_2_, *O*_3_, and *CO*) with COVID-19 daily new cases and COVID-19 daily new deaths. At a moving average of 14 days (lag0-14 days), *PM*_2.5_, *PM*_10_, and *NO*_2_ were significantly associated with increased risk of COVID-19 daily new cases, whereas all the ambient air pollutants *PM*_2.5_, *PM*_10_, *SO*_2_, *NO*_2_, *O*_3_, and *CO* were significantly associated with COVID-19 daily new deaths. Similarly, At a moving average of 21 days (lag0-21 days), *PM*_10_, *SO*_2_, and *NO*_2_ were significantly associated with increased risk of COVID-19 daily new cases. In contrast, all the ambient air pollutants *PM*_2.5_, *PM*_10_, *SO*_2_, *NO*_2_, *O*_3_, and *CO* were significantly associated with COVID-19 daily new deaths. It suggests that at a moving average of 14 days (lag0-14 days), *PM*_2.5_, *PM*_10_, and *NO*_2_ were significantly associated with increased risk of both COVID-19 daily new cases and COVID-19 daily new deaths. Likewise, at a moving average of 21 days (lag0-21 days), *PM*_10_, *SO*_2_, and *NO*_2_ were significantly associated with increased risk of both COVID-19 daily new cases and COVID-19 daily new deaths. On the other hand, the effects of ambient air pollution on COVID-19 daily new cases and COVID-19 daily new deaths were not statistically significant at a moving average of 7 days (lag0-7 days). It is of great significance to state that in sensitivity analysis after adjusting for population mobility in terms of lockdowns, the effects of ambient air pollution on COVID-19 daily new cases and COVID-19 daily new deaths became more prominent and robust.

Many previous time-series studies demonstrated that the ambient air pollution is closely related to respiratory diseases (Ciencewicki and Jaspers, 2007; Horne et al., 2018; Mehta et al., 2013; Phosri et al., 2019; Tramuto et al., 2011; Xie et al., 2019; Zhang et al., 2015). However, very few studies have examined the association of ambient air pollution with COVID-19 daily new cases and COVID-19 daily new deaths. A time-series study in China reported significant positive associations of *PM*_2.5_, *PM*_10_, *NO*_2_, *O*_3_, and *CO* with COVID-19 daily new cases at lag0-7 and lag0-14 (Zhu et al., 2020). Though, *SO*_2_ was negatively associated with the COVID-19 daily new cases. In contrast, we observed that *SO*_2_ was positively associated with COVID-19 daily new cases. The underlying mechanism of *SO*_2_ and COVID-19 daily new cases needs to be further investigated. A multi-city time-series study conducted in China also observed that *PM*_2.5_ and *PM*_10_ were significantly associated with the increased risk of daily confirmed COVID-19 cases (Wang et al., 2020). In line with our study, a time-series and machine learning study in New York City also confirmed that *PM*_2.5_ is associated with a daily COVID-19 infection incidence at lag0-14 (Mirri et al., 2021).

Our study also established the strong associations between ambient air pollution and COVID-19 daily new deaths. Earlier studies have shown that *PM*_2.5_ is positively associated with COVID-19 deaths (Frontera et al., 2020; Mele and Magazzino, 2021; X. Wu et al., 2020; Zhou et al., 2020; Zhu et al., 2020). Our findings fall in line with these papers as we have revealed that *PM*_2.5_ is positively associated with COVID-19 daily new deaths at lag0-14 and lag0-21. A study by Jiang and Xu (2021) using Poisson regression reported that *PM*_2.5_, *PM*_10_, *SO*_2_, and *CO* were strongly associated with COVID-19 deaths. Among them, *PM*_2.5_ was the single variable that showed a positive association with COVID-19 deaths. Contrary to this, our findings revealed that *PM*_2.5_, *PM*_10_, *NO*_2_, *O*_3_, and *CO* were positively associated with daily new COVID-19 deaths at lag0-14 and lag0-21. The apparent difference between their study and ours is that we used a time-series study design to examine the association between air pollution and COVID-19 daily new deaths. Consistent with the previous study by Wang et al. (2020), after controlling for the population mobility, the associations of *PM*_2.5_, *PM*_10_, *NO*_2_, *O*_3_, and *CO* with COVID-19 daily new cases and COVID-19 daily new deaths become more prominent. It is worthwhile to mention that *O*_3_ at all lags and *CO* at lag0-14 and lag0-21 were significantly associated with COVID-19 daily new deaths only.

There are some limitations to this study. First, the causal effects of air pollution on COVID-19 daily new cases and COVID-19 daily new deaths were not established as we only concentrated on the associations of air pollution with COVID-19 daily new cases and COVID-19 daily new deaths. Second, other than the COVID-19 daily new cases and COVID-19 daily new deaths, the demographic data of COVID-19 were unavailable, so we could not carry sub-population analyses.

## Conclusions

Our study concludes that there are statistically significant associations of ambient air pollutants *PM*_2.5_, *PM*_10_, *SO*_2_, and *NO*_2_ with COVID-19 daily new cases and COVID-19 daily new deaths. *O*_3_ and *CO* were significantly associated with COVID-19 daily new deaths only. Besides, in sensitivity analysis after controlling the population mobility in terms of lockdowns these associations became more prominent. These findings suggest that governments need to give greater considerations to regions with higher concentrations of *PM*_2.5_, *PM*_10_, *SO*_2_, *NO*_2_, *O*_3_, and *CO*, since these areas may experience a more serious COVID-19 pandemic or, in general, any respiratory disease.

## Data Availability

Publicly available data were used in the study.

## Electronic Supplementatry Materials

**Table S1:**
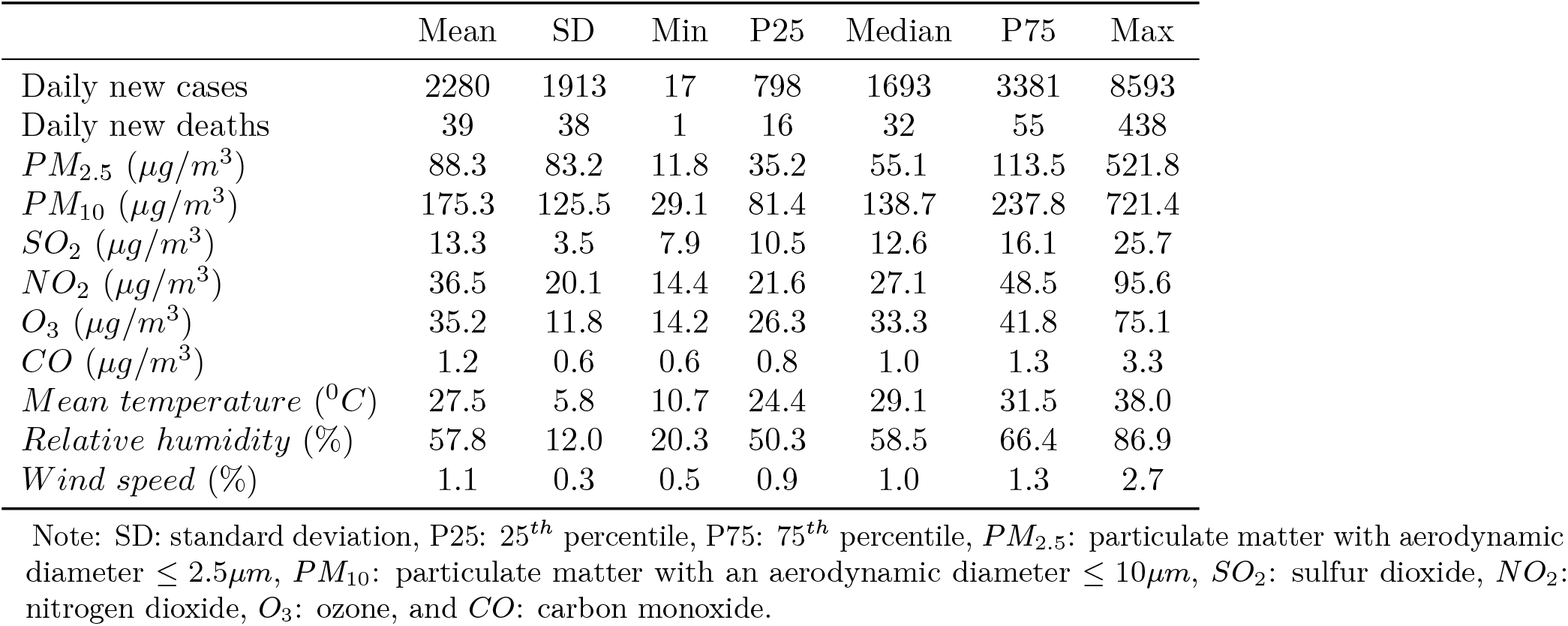
Summary statistics for daily new cases and daily new deaths of COVID-19, air pollutants, and meteorological variables in National Capital Territory of Delhi, India during the study period (1 April 2020 through 31 December 2021).

**Table S2:**
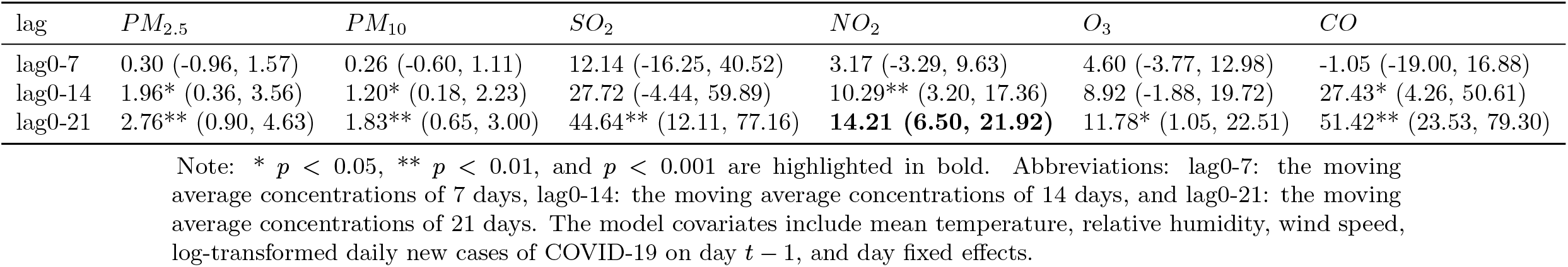
Percent change (means and 95% confidence intervals) in COVID-19 daily new cases associated with a 10*— µg/m*^3^ increase in *PM*_2.5_, *PM*_10_, *SO*_2_, *NO*_2_, and *O*_3_ and 1−*µg/m*^3^ increase in *CO* using different lag days in single pollutant models after controlling the population mobility.

**Table S3:** Percent change (means and 95% confidence intervals) in COVID-19 daily new deaths associated with a 10 *—µg/m*^3^ increase in *PM*_2.5_, *PM*_10_, *SO*_2_, *NO*_2_, and *O*_3_ and 1*− µg/m*^3^ increase in *CO* using different lag days in single pollutant models after controlling the population mobility.

| lag | $PM_{2.5}$ | $PM_{10}$ | $SO_2$ | $NO_2$ | $O_3$ | $CO$ |
| --- | --- | --- | --- | --- | --- | --- |
| lag0-7 | -0.30 (-2.08, 1.56) | 0.22 (-0.99, 1.43) | 39.02 (-2.11, 80.16) | 9.33* (1.07, 17.58) | **27.03 (14.23, 39.83)** | -8.81 (-35.54, 17.92) |
| lag0-14 | 3.63** (1.33, 5.94) | **2.59 (1.15, 4.03)** | **105.71 (61.42, 150.01)** | **21.63 (11.95, 31.30)** | **48.83 (31.42, 66.23)** | **64.04 (31.11, 96.97)** |
| lag0-21 | **7.48 (4.59, 10.36)** | **4.76 (3.00, 6.53)** | **132.07 (81.98, 182.16)** | **29.66 (18.14, 41.17)** | **69.05 (48.55, 89.55)** | **146.89 (103.41, 190.37)** |

Note: \* $p < 0.05$ , \*\* $p < 0.01$ , and $p < 0.001$ are highlighted in bold. Abbreviations: lag0-7: the moving average concentrations of 7 days, lag0-14: the moving average concentrations of 14 days, and lag0-21: the moving average concentrations of 21 days. The model covariates include mean temperature, relative humidity, wind speed, log-transformed daily new cases of COVID-19 on day $t - 1$ , and day fixed effects.

## Notes

### Competing Interest Statement

The authors have declared no competing interest.

### Funding Statement

None

